# Vascular brain pathology is more important than neurodegeneration in pathogenesis of pre-stroke cognitive impairment

**DOI:** 10.1101/2020.06.12.20129106

**Authors:** T Schellhorn, M Zucknick, T Askim, R Munthe-Kaas, H Ihle-Hansen, YM Seljeseth, AB Knapskog, H Næss, H Ellekjær, P Thingstad, T Bruun Wyller, I Saltvedt, MK Beyer

## Abstract

**Introduction:** To better understand the development of post-stroke cognitive impairment,we explored the association between pre-stroke neuroimaging features and pre-stroke cognitive impairment and investigated possible gender differences. Few previous studies on this topic have been performed.

**Methods:** In this large prospective longitudinal multicenter brain-MRI cohort study, patients admitted to five stroke units at five different Norwegian hospitals were recruited as part of the Norwegian cognitive impairment after stroke study. Visual radiological assessment of small vessel disease and neurodegenerative changes were performed on brain MRI from 410 patients. Pre-stroke cognition was assessed using the Global Deterioration Scale.

**Results:** At least one pathological marker was found in 68% of the patients. The mean age (SD) of the patients with no pathological changes other than the acute stroke, was 70 (± 12.9) and 75 (± 10.2) years for those with pathological scans (p ≤ 0.001). Men were more likely to have at least one pathological brain MRI finding, lacunes, or pathological medial temporal lobe atrophy. The highest percentage of patients with a pathological pre-stroke GDS were found in the “cerebrovascular pathology” group (37.5%) and in the “mixed pathology”-group (44%). In these groups, both men and women had an increased risk of impaired pre-stroke cognition.

**Conclusion:** The majority of patients showed preexisting structural brain pathology. Cerebrovascular pathology was the dominating imaging finding associated with cognitive impairment, thus indicating that the pathogenesis of pre-stroke cognitive impairment might be driven more by small vessel disease (SVD) than neurodegenerative changes. Gender differences exists, with less pathology in women.

## Introduction

Post-stroke dementia (PSD) occurs in about 20 % of patients with a history of stroke [1]. The etiology of PSD is not yet fully understood, but brain resilience seems to be a major determining factor for whether patients develop PSD. One of the risk factors for PSD is pre-stroke cognitive status [2] which also has been shown to have a negative effect on survival in PSD patients[3]. Studies looking at risk factors for pre-stroke cognitive impairment are scarce compared to studies of causes for PSD. One large study found that older age at onset, greater prevalence of atrial fibrillation, history of stroke, heart failure and premorbid use of anticoagulants and anti- hypertensive medication were associated with cognitive impairment prior to stroke [4]. Few studies of brain changes associated with pre-stroke cognitive impairment exist. One study of 78 stroke patients found that cerebral atrophy was associated with prestroke cognitive impairment[5].

Women who survive a stroke have a poorer functional outcome, a higher need for home nursing, and show higher rates of recurrent stroke [6]. Additionally, the female sex is more strongly associated with pre-stroke dementia [7], with most of these circumstances being attributed to the fact that women are older than men at the time of their first stroke [6]. Whether female sex is associated with pre-stroke cognitive impairment, and risk factors for this has to our knowledge not yet been studied.

Brain magnetic resonance imaging (MRI) is a valuable tool for characterizing structural brain changes. Pre-stroke chronic brain changes are important predictors of PSD, lowering the threshold for the development of PSD [8]. Measuring the pre-stroke structural disease burden and it’s relationship to pre-stroke cognitive impairment might help assist the prediction of development of cognitive decline after an acute stroke and provide increased insight into pathogenetic mechanisms for post-stroke cognitive impairment. Lacunes, white matter hyperintensities (WMH) and cerebral microbleeds are typical MRI signs of small vessel disease (SVD), which is believed to be the main etiology for about 20% of the cases of ischemic stroke [9]. Pre-stroke SVD is gaining increasing attention as an important predictor of cognitive decline and dementia [10] [11].This is especially true for vascular dementia [12], but also for Alzheimer disease [13]. Typical imaging features of neurodegeneration are medial temporal lobe atrophy (MTA), ventricular enlargement, and posterior atrophy [14]. MTA constitute an early sign of Alzheimer’s disease [15] and is accelerated in patients with mild cognitive impairment [16].

Neurodegeneration is frequent in the older population and commonly coexists with cerebrovascular disease [9]. Markers of SVD are found in brain imaging in healthy elderly people, with lacunes occurring in 20% and WMH in up to 95% [17, 18]. Consequently, cut-offs must be used to distinguish normal aging from pathological changes [14].

Previous studies have focused on WMH as an imaging marker of SVD, and MTA as an imaging marker of neurodegeneration [2, 19, 20]. Studies that investigate established imaging markers for both SVD and neurodegeneration in one population are lacking. Little is known about the role of SVD, neurodegeneration and gender in the pathogenesis of pre-stroke cognititive changes. It is therefore important to study pre-stroke structural brain changes combined with measures of pre-stroke cognition.

In this study we wanted to characterize the burden of preexisting brain pathologies in a cohort of patients admitted to the hospital with acute stroke.

The objectives of this study were to a) To characterize pre-stroke neurodegenerative and vascular disease burden found on brain MRI, b) To describe the association between pre-stroke neuroimaging features and pre-stroke cognitive impairment, and c) investigate possible gender differences in the risk of prestroke cognitive impairment.

## Methods

The Norwegian cognitive impairment after stroke (Nor-COAST) study is a prospective longitudinal multi-center cohort study [21]. Patients admitted to the stroke units at the following Norwegian hospitals were recruited; Ålesund hospital, St. Olav University Hospital, Haukeland University Hospital, Oslo University Hospital, and Bærum Clinic of Vestre Viken Hospital. Patient recruitment in Nor-COAST started in May 2015 and was completed in March 2017. The study protocol has been approved by the regional committees for medical and health research, REK Nord (REK number: 2015/171), and registered in the clinicaltrials.gov (NCT02650531).

The regional committees for medical and health research, REK Nord, have approved this sub study (REK number: 2016/2306). Participation in the study is voluntary. The participants gave their informed written consent before inclusion in accordance with the Declaration of Helsinki. When a person was unable to give their consent, informed written consent for participation was given by a family proxy. Participants were asked specifically if they would be interested to take a brain MRI-scan as part of the study.

### Population

Inclusion criteria were: 1)Hospitalization with acute ischemic or haemorrhagic stroke hospitalized within one week after onset of symptoms. Acute stroke, diagnosed according to the World Health Organization (WHO) criteria 2) age over 18 years, 3) fluent in Scandinavian language. Exclusion criteria: 1) not treated in the participating stroke units 2) symptoms explained by other disorders than ischemic brain infarcts or intra-cerebral hemorrhages. 3) expected survival less than 3 months after stroke. Inclusion criteria for MRI: 1)Patient included in Nor-COAST, 2) modified Rankin scale <5 before the stroke, 4) able to cooperate during MRI. Exclusion criteria for MRI: 1)Severe functional impairment making MRI impossible to perform, 2) medical contra-indications for MRI like claustrophobia or pacemaker 3) patient decline participation in MRI.

### MRI acquisition

A study-specific brain MRI was taken in the acute/subacute phase of the stroke (i.e. 2-7 days after symptom onset). Brain scans were acquired at five different sites, but in the same MR-scanner on each site (GE Discovery MR750, 3T; Siemens Biograph_mMR, 3T; Philips Achieva dStream, 1.5T; Philips Achieva, 1.5T; Siemens Prisma, 3T). The study protocol consisted of a 3D-T1 weighted, axial T2, 3D-Fluid attenuated inversion recovery (FLAIR), diffusion weighted imaging (DWI) and susceptibility weighted imaging (SWI). Details about the MRI protocol can be found in table 1 of the Supplementary material. In addition, patients had a clinical CT or MRI taken at admittance to the hospital in the acute phase. Some of these MRIs were also available to us at the time for visual analysis (see Figure 1). Remaining clinical brain scans are not yet ready for analyses. No detailed information was obtained on patients who were not referred to a studyspecific MRI.

**Table 1.**
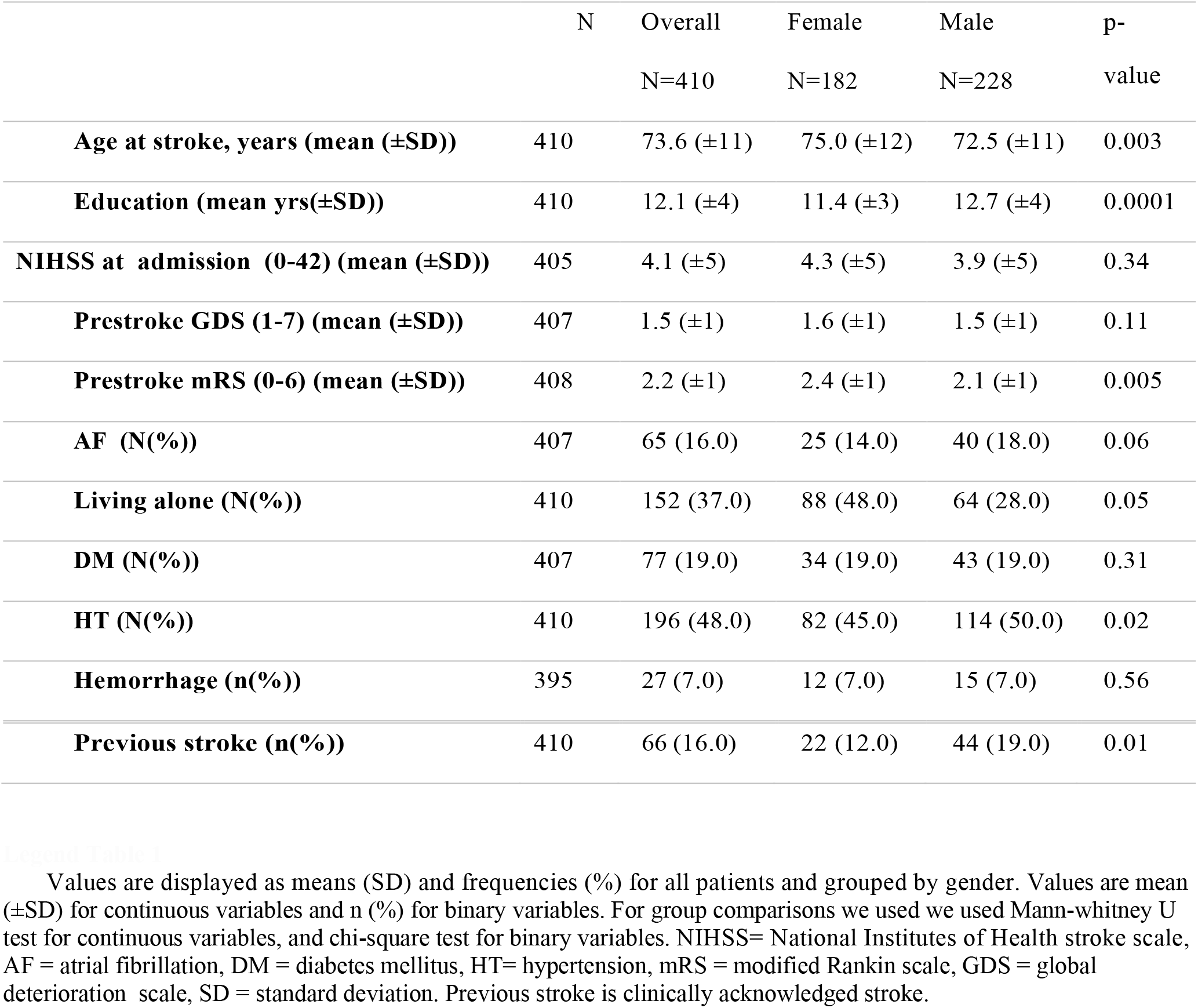
Baseline characteristics of the included patients

|  | N | Overall | Female | Male | p-value |
| --- | --- | --- | --- | --- | --- |
|  |  | N=410 | N=182 | N=228 |  |
| **Age at stroke, years (mean (±SD))** | 410 | 73.6 (±11) | 75.0 (±12) | 72.5 (±11) | 0.003 |
| **Education (mean yrs(±SD))** | 410 | 12.1 (±4) | 11.4 (±3) | 12.7 (±4) | 0.0001 |
| **NIHSS at admission (0-42) (mean (±SD))** | 405 | 4.1 (±5) | 4.3 (±5) | 3.9 (±5) | 0.34 |
| **Prestroke GDS (1-7) (mean (±SD))** | 407 | 1.5 (±1) | 1.6 (±1) | 1.5 (±1) | 0.11 |
| **Prestroke mRS (0-6) (mean (±SD))** | 408 | 2.2 (±1) | 2.4 (±1) | 2.1 (±1) | 0.005 |
| **AF (N(%))** | 407 | 65 (16.0) | 25 (14.0) | 40 (18.0) | 0.06 |
| **Living alone (N(%))** | 410 | 152 (37.0) | 88 (48.0) | 64 (28.0) | 0.05 |
| **DM (N(%))** | 407 | 77 (19.0) | 34 (19.0) | 43 (19.0) | 0.31 |
| **HT (N(%))** | 410 | 196 (48.0) | 82 (45.0) | 114 (50.0) | 0.02 |
| **Hemorrhage (n(%))** | 395 | 27 (7.0) | 12 (7.0) | 15 (7.0) | 0.56 |
| **Previous stroke (n(%))** | 410 | 66 (16.0) | 22 (12.0) | 44 (19.0) | 0.01 |

**Figure 1.**
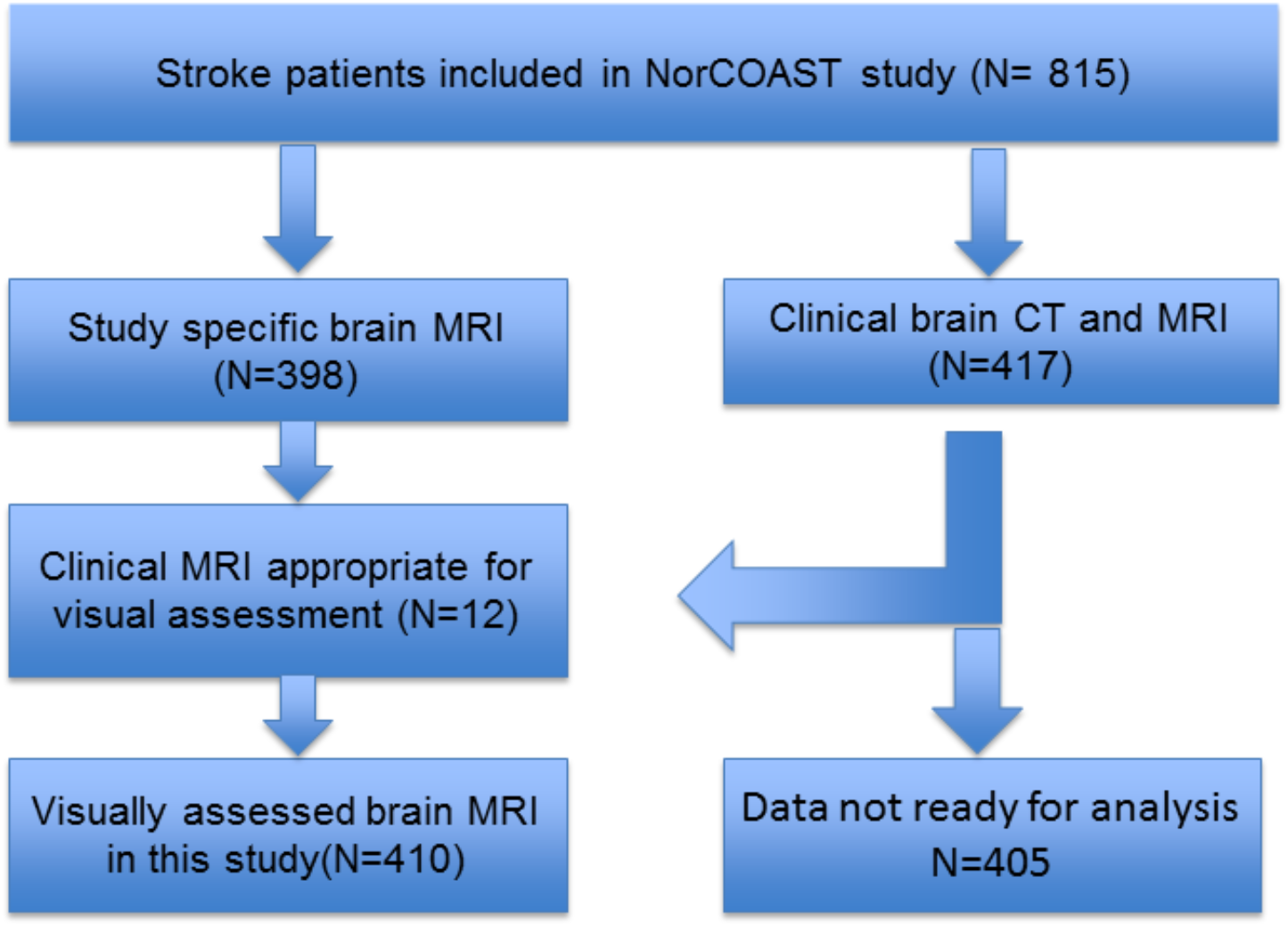
Flow chart of patient imaging in the NorCOAST study

### Image analysis

In this study we only evaluated brain changes not related to the acute stroke. We analyzed our images according to validated visual rating scales for imaging markers of neurodegenerative disease [14]. SVD features were rated according to Standards for Reporting Vascular Changes on Neuroimaging (STRIVE) recommendation [9]. The extent of white matter hyperintensities of presumed vascular origin (in the following abbreviated as WMH) was classified according to the widely used Fazekas scale [22], and categorized as ‘normal’ with a Fazekas grade of 1. Fazekas grade 2 is considered normal in patients 71 years or older, whereas Fazekas grade 3 was always regarded as pathological [23].

Lacunes of presumed vascular origin (in the following abbreviated as lacunes), are defined as cystic lesions with a diameter between 3 and 15 mm, surrounded by hyperintense T2-FLAIR signal. Lacunes were always regarded as pathological [24].

Microbleeds are round/ovoid lesions with low signal on SWI ≤ 10mm. For brain microbleeds we dichotomized the BOMBS scale so that patients were registered either to have or not to have microbleeds. [25] To minimize false positive ratings, the existence of more than one hypointense lesions on SWI was regarded as presence of microbleeds [26]. The presence of microbleeds was always regarded as pathological.

Medial temporal lobe atrophy was assessed according to the established MTA scale [27]. A mean MTA of both sides ≥ 1.5 was considered pathological under the age of 75, a value ≥ 2 below the age of 85, and a value of ≥ 2.5 below 95 years, as recommended by Ferreira et al. [14].

Posterior atrophy was assessed according to the posterior atrophy (PA) scale [28]. A value of ≥ 2 was considered pathological in patients below 95 years [14].

Ventricular enlargement was measured by the Evans index (EI)[29]. Sex and age dependent reference values for the Evans index was taken from Brix et al.[30].

Patients with similar combinations of imaging markers were grouped together. The groups were clustered into; 1. A “Neurodegeneration” group, with pathological scores only for MTA, PA or Evans index; 2. A “cerebrovascular pathology” group, with pathological scores for WMH, lacunes or microbleeds; 3. A “mixed pathology” group, with patients with pathological scores for imaging markers from both categories 1 and 2; 4. A “Normal” group, with no pathological scores on visual rating of brain MRI.

### Cognitive assessment and cerebrovascular risk factors

Pre-stroke cognitive impairment was assessed by interview of relatives or caregivers, using the Global Deterioration Scale (GDS)[31]. Using GDS we assessed the global cognitive function of the patients during the time before the acute stroke. There are seven stages in the GDS with increasing cognitive impairment from stage one to seven. Stage one is no cognitive impairment and in stage seven, the patients have severe dementia. Patients with a GDS score of two are categorized as very mild cognitive/subjective cognitive impairment [31]. Consequently GDS 2 represents the first stage in a pathological continuum. Since GDS 2 is considered the first stage of pathological decline we rated it as pathological. [32].Patients with GDS 1 were scored as not having cognitive impairment. We used the GDS score to analyze the association between imaging marker groups and cognitive function before stroke. The modified Rankin Scale (mRS) was used to measure general functioning and the degree of disability in daily activities, with scores ranging from zero (independent) to six (dead) [33].

The following criteria were used in defining clinical diagnoses and risk factors in the Nor-COAST study: Atrial fibrillation (AF) was defined as patients with a pathological ECG recording documenting this heart rythm, past or present. Hypertension (HT) was noted in patients using anti-hypertensive drugs. Hypercholesterolemia was defined by total cholesterol ≥ 6.2 mmol/L or LDL ≥ 4.1 mmol/L. Diabetes mellitus (DM) was registered when evident in medical records, when the patient used antidiabetic medication, or by a HbA1c ≥ 6.5%. Previous stroke was registered when medical records showed a history of stroke. Stroke severity was assessed by the National Institute of Health Stroke Scale (NIHSS)[34].

### Statistical analysis

For group comparisons we used we used Mann-whitney U test for continuous variables, and the chi-square test was used for binary variables.

The Odds ratio (OR) of having a pathological GDS was calculated with univariate logistic regression analysis for age above mean versus age below mean, gender, pathological imaging group,silent infarcts, education, hypertension, artrial fibrillation and clinically acknowledged previous stroke. Education groups were dichotomized into patients having university level education defined as ≥ 15 years of education and patients having < 15 years of education. Silent infarct was defined as patients with visible MRI changes of either lacunes or cortical/subcortical infarct, but no previous clinical infarct.

The relationship between gender and pathological imaging findings was modeled with logistic regression, using male gender as the reference group.

Since age represents an important potential confounding variable it was explicitly adjusted for in the logistic regression models. This is also true for models where explanatory variables ie brain imaging markers, already were age-adjusted.

The relationship between the imaging groups and the existence of a pathological GDS was modeled with a multiple logistic regression with “Normal” set as the reference group. In this model we additionally evaluated the effect of imaging groups plus gender, previous clinical stroke and gender-by-imaging group interaction as confounding variables. The difference between the “mixed”- and “cerebrovascular”-group was explored with a logistic regression model with “Cerebrovascular” set as reference and the effect of gender and gender-by-imaging group interaction was evaluated.

All effect sizes are represented by ORs with 95% confidence intervals (CI), both unadjusted and adjusted for gender.

The logistic regressions were done with help of the “statsmodels” package version “0.10.1” for Python version “3.6.7”. Two-sided p-values below 0.05 were considered to indicate statistical significance.

To test interrater agreement of visual assessment, two pilots of 20 scans, and later 9 new scans were selected and scored by 2 experienced neuroradiologists independently (TS, 12 yrs experience; MKB 15 yrs experience). Causes for diverging ratings were identified and resolved in a consensus meeting. Finally, 30 scans were independently rated by both neuroradiologists. The intraclass correlation coefficient (ICC, model “two-way” and type “agreement”) and its 95% CI were calculated as a measure of the interrater agreement with the “irr” package version 0.84.1 for R [35] version 3.4.1 [36]. The same 30 scans were rated twice by one rater (TS) and intra-rater reliability was repeatedly calculated. Cohens kappa and its 95% confidence interval was calculated as a measure of interrater agreement for the binary imaging scores with help of the “irr” package version 0.84.1 for R [35].

## Results

### Study Population

A total of 815 patients were enrolled in the Nor-COAST study. Study specific MRI-scans were performed in 398 (48.8.%) participants. The remaining 417 (51.2%) patients were not part of the MRI-study due to severe functional impairment, medical contra-indications, or because the patient declined participation in the MRI sub-study. 12 of the clinical MRI scans were used in this study.

The clinical and demographic characteristics and cerebrovascular risk factors are summarized in Table 1. The mean age (SD) was 73.6 (±11) years. The mean National Institute of Health Stroke Scale (NIHSS) score at admission (SD) was 4.1(±5). Females were significantly older than males (75 vs 72.5yrs), had less education (11.4 vs 12.7 yrs), and a higher pre-stroke mRS (2.4 vs. 2.1). Clinically acknowledged previous stroke was found in 66 (16.1%) of the patients. Previous infarcts not clinically acknowledged were found in 139 (34%) of the patients. Significantly more men had suffered a previous clinically acknowledged stroke (19% vs 12%) and had hypertension (50% vs 45%).

*<u>Table 1 Population characteristics</u>*

### Imaging findings

The visual scoring of chronic brain changes on the MRI-scans showed at least one pathological finding in 278 (68%) of the patients. The mean age (SD) of the patients with normal brain was 70 (± 12.9) and 75 (± 10.2) years for those with pathological scans (p ≤ 0.001)

*<u>Table 2 - Absolute and relative frequencies of imaging findings for the whole population and grouped by gender</u>*.

Mean Fazekas score (SD) was 1.8 (±0.9), and a pathological WMH-score was present in 154 (38%). Visible evidence of infarctions other than the index stroke was found in 78 patients (20%). Lacunes were identified in 142 (35%) and microbleeds were found in 76 (19%) of the patients.

**Table 2.** Pathological MR-imaging findings. This table shows the number of patients who have different types of pathological MRI scores in our cohort, and the gender distribution of pathological scores.

|  | Total n/N (%) | Female<br>n/N (%) | Male n/N<br>(%) | Unadjusted<br>OR (†)<br>(95% CI) | p-value | Adjusted OR<br>(††)<br>(95% CI) | p-value |
| --- | --- | --- | --- | --- | --- | --- | --- |
| At least 1 path. | 278/410 (68) | 108/182<br>(59) | 170/228<br>(75) | 0.5<br>(0.33-0.76) | 0.001* | 0.43<br>(0.28-0.66) | 0.0002* |
| WMH | 154/410<br>(38) | 74/182<br>(41) | 80/228<br>(35) | 1.3<br>(0.9-1.9) | 0.25 | 1.13<br>(0.7- 1.7) | 0.57 |
| Old infarction | 78/396<br>(20) | 28/175<br>(16) | 50/221<br>(23) | 0.65<br>(0.4-1.1) | 0.10 | 0.62<br>(0.4-1.0) | 0.07 |
| Lacunes | 142/410<br>(35) | 51/182<br>(28) | 91/228<br>(40) | 0.6<br>(0.4-0.9) | 0.01* | 0.53<br>(0.3-0.8) | 0.003* |
| MicroB | 76/410<br>(19) | 33/182<br>(18) | 43/228<br>(19) | 0.95<br>(0.6-1.6) | 0.85 | 0.93<br>(0.6-1.5) | 0.78 |
| MTA | 125/410<br>(30) | 34/182<br>(19) | 91/228<br>(40) | 0.35<br>(0.2-0.6) | 0.001* | 0.35<br>(0.2-0.6) | 0.001* |
| PA | 44/410<br>(11) | 17/182<br>(9) | 27/228<br>(12) | 0.77<br>(0.4-1.5) | 0.42 | 0.57<br>(0.3-1.1) | 0.11 |
| Evans index | 17/410<br>(4) | 6/182<br>(3) | 11/228<br>(5) | 0.67<br>(0.2-1.9) | 0.44 | 0.7<br>(0.3-1.9) | 0.49 |

A pathological MTA-score was present in 125 (30%) and a pathological PA score in 44 (11%) of all patients, while only 17 (4%) had pathologically enlarged ventricles.

Significantly fewer women than men showed at least one pathological finding (108 (59%) vs.

170 (75%)). Compared to men, fewer women had lacunes (51 (28%) vs. 91 (40%)) and medial temporal lobe atrophy (34 (19%) vs. 91 (40%)). These differences remained when adjusted for age (see Table 2).

### Cognitive assessment and cerebrovascular risk factors

As shown in Table 3, 100 of 276 (36%) patients with baseline pathological MRI also had a pathological GDS score, while 25 of 143 (18.9%) with normal MRI had a pathological GDS score.

**Table 3.** Gender distribution of imaging pathology and prestroke cognitive impairment

| Pathological Imaging group (N(%)) | GDS 1 N(%) |  | GDS ≥2 N(%) |  | Unadjusted Odds ratio (CI) (†) | p-value | Adjusted Odds ratio (CI) (††) | p-value |
| --- | --- | --- | --- | --- | --- | --- | --- | --- |
|  | 282(69) |  | 125(31) |  |  |  |  |  |
|  | Male | Female | Male | Female |  |  |  |  |
|  | 161(57) | 121(43) | 65(52) | 60(48) |  |  |  |  |
| Normal (131(32)) | 46(43) | 60(57) | 12(48) | 13(52) | 1 |  | 1 |  |
| Cerebrovascular pathology (120(29)) | 40(53) | 35(47) | 18(40) | 27(60) | 3.0 (1.8-5.1) | 0.001* | 3.1 (1.9- 5.3) | 0.001* |
| Mixed pathology(99(24)) | 40(73) | 15(27) | 27(61) | 17(39) | 3.7 (2.3- 6.1) | 0.001* | 4.2(2.6-7.0) | 0.001* |
| Neurodegeneration(57(14)) | 35(76) | 11(24) | 8(73) | 3(27) | 1.1 (0.5-2.3) | 0.8 | 1.3 (0.6-2.7) | 0.5 |

Using univariate logistic regression analysis we found that the OR(95% CI) for a pathological GDS for old age was 3.5 (2.21-5.49) compared to the patients with age below mean. For patients with clinically acknowledged previous infarct OR (95% CI) was 2.9 (1.7- 5.0) compared to patients who had not suffered a clinical stroke previously. For the cerebrovascular pathology group 2.54 (1.44-4.5), for the mixed pathology group 3.39 (1.88- 6.11) and for the neurodegeneration group 1 (0.46-2.23). The OR (95% CI) for a pathological GDS in patients with higher education was 0.26 (0.15-0.45) which was significantly reduced versus patients with a low education level (p< 0.001).The OR (95% CI) for a pathological GDS for gender, atrial fibrillation, silent infarcts and hypertension did not reach statistical significance.

The highest percentage of patients with a pathological pre-stroke GDS were found in the “cerebrovascular pathology”-group with 37.5%, and in the “mixed pathology”-group with 44 %, while the lowest prevalence of a pathological pre-stroke GDS was found in the “neurodegeneration”-group with 19%.

In the multiple logistic regression model the OR (95% CI) for having a pathological GDS compared to the “Normal”-group was 3.0 (1.8-5.1) for the “cerebrovascular pathology”-group and 3.7(2.3-6.1) for the “mixed pathology”-group and 1.1(0.5-2.3) for the “neurodegeneration”-group (see table 3). Correcting for gender, previous clinical stroke and the interaction gender by imaging group did not change the results significantly. Correcting for education and imaging group did not significantly change the OR for having a pathological GDS. Patients in the “mixed pathology” - and the “cerebrovascular pathology” groups had significantly higher OR of having a pathological pre-stroke GDS than patients in the “normal” group.

The OR for having a pathological GDS (95% CI) in the “mixed pathology”-group is 1.3 (CI 0.92-1.95) compared to the “cerebrovascular pathology”-group (p= 0.13).

*<u>Table 3 Patients grouped according to imaging pathology and prestroke cognitive impairment (GDS)</u>*

### Reliability

The ICC (95% CI) indicated excellent reliability for the Fazekas-scale 0.9 (0.84-0.96), good reliability for the EI 0.9 (0.82-0.96) and MTA-scale 0.8 (0.59-0.0.89), with the PA scale showing moderate reliability 0.5 (0.2-0.74).

Cohens kappa (95% CI) showed fair/moderate agreement for the rating of lacunes 0.4(-.23- 0.35), substantial agreement for DWI-lesions 0.6 (0.28-0.91), and almost perfect agreement for microbleeds 0.9 (0.67-1). For macrobleeds we found perfect agreement.

The ICC (95% CI) for the intra-rater agreement indicated excellent agreement for the Fazekas-scale 0.9 (0.7-0.93), MTA-scale 0.8 (0.64-0.91), and Evans-index 0.9 (0.75-0.96). The agreement for the Koedam scale was poor 0.4 (0.01-0.62). Cohen’s kappa indicated moderate intra-rater agreement for lacunes 0.5 (−0.22-0.38), slight agreement for microbleeds 0.2 (−0.24- 0.55), and perfect agreement for macrobleeds.

## Discussion

Nor-COAST is a large multicenter brain-MRI study of a prospectively sampled Norwegian stroke population on patients with acute stroke

Apart from the acute stroke lesion, we found at least one pathological marker on the MRI of 68% of the patients. Patients with a normal appearing brain before the acute stroke were significantly younger than the patients with pathological brain scans.

The two most common imaging findings were WMH and lacunes while MTA was the third most common pathology. Women were older than men, but had fewer pathological markers including MTA and lacunes. More men had hypertension and a previous clinically acknowledged stroke.

Pre-stroke cognitive impairment was found in 36% of the patients with a pathological MRI, while only 19% of the patients with a normal appearing MRI had a pathological GDS score. The highest percentages of patients with a pathological pre-stroke GDS were found in the “cerebrovascular pathology”-group and “mixed pathology” - and the “cerebrovascular pathology” had significantly higher OR of having a pre-stroke cognitive impairment than patients in the “normal” group, while the OR of “mixed pathology” group compared to “cerebrovascular pathology” group were not significantly different.

The mean NIHSS was 4.1 at admission. Despite suffering minor to moderate strokes, we found pre-stroke imaging pathology to be present in more than two thirds of all patients in their baseline scans which is a substantial pre-stroke disease burden. More than half of all patients showed markers of SVD, and one in three had neurodegenerative changes exceeding what is normal for their age according to published cutoff values.

WMH was the most prevalent imaging marker of SVD, and MTA the most prevalent imaging marker of neurodegeneration. Consistent with our results, previous studies have shown that imaging markers of SVD and MTA are frequently present in patients with stroke. Takahashi found MTA grade 2-3 (on a modified Scheltens scale) in 75% of all cases in a study of 69 stroke patients within 14 days after stroke [37]. In the “GRECogVASC Study”, Puy found a MTA score of 2.5 or greater in 9.8% of 356 stroke patients (mean age 64 yrs.) six months after the acute event [38]. In studies of patients aged 90 years and older cerebrovascular disease is the most prevalent non Alzheimer age associated pathology[39].

Microbleeds were present in 22% of the patients in the “GRECogVASC Study”; a four percent higher rate than what was found in our study. In the population-based “AGES”-study of Sveinbjornsdottir and colleagues, the prevalence of microbleeds was 11 %, based on the brain MRI scans of 1962 patients with a mean age of 76 years [40]. Akhtar and collegues found microbleeds in 22% of all patients in an acute stroke population [41]. These studies show that the prevalence of microbleeds is about twice as high compared to the healthy ageing population. Mixed-location cerebral microbleeds have been associated with cognitive impairment and dementia in the presence of cerebrovascular diseases [42], and may therefore be an important predictor of cognitive impairment after stroke. Since this is a study of stroke patients it is not suprising to have a high prevalence of markers of SVD.

Many previous studies have looked into the association of single risk factors or single brain markers and eg post-stroke dementia. Few have grouped brain markers in categories, even though it is known that patients with mixed pathologies of neurodegenerative and cerebrovascular changes is the largest group [43]. In our study, imaging markers of cerebrovascular disease or a combination of cerebrovascular disease and neurodegeneration showed a stronger association with a pathological pre-stroke GDS than markers of neurodegeneration alone. Patients without any pathological imaging markers or with imaging markers of neurodegeneration showed a weaker association with a pathological pre-stroke GDS, indicating preserved cognition. Clinically verified previous stroke did not affect this association significantly although there was a significantly increased OR for pathological GDS in the univariate analysis of patients with previous clinical stroke.

The odds of having a pathological pre-stroke GDS was higher in patients in the mixed imaging group compared to the other pathological imaging groups. It is not surprising that the “mixed” group have higher odds for pre-stroke cognitive impairment than the “cerebrovascular”-group, indicating an additive effect of neurodegeneration and cerebrovascular pathology. This suggests that the cerebrovascular changes alone, or in combination with neurodegenerative changes affects pre-stroke cognition the most. This is in line with a study focusing on risk factors for developing dementia which showed that additional non-Alzheimer pathology reduced the odds of being resilient [44]. Robinson et al also found that in The 90+ Study, resilient patients had less cerebrovascular disease [45]. This is in conflict with the hypothesis that most of the cognitive decline in a stroke population is caused by coexisting neurodegenerative changes, as suggested by Jokinen et al. [46].

Another explanation could be that the neurodegenerative component in the “mixed”-group is linked to SVD. This is an association that has already been described for WMH and MTA [47] [48], suggesting that the neurodegenerative changes in these patients are secondary to cerebrovascular disease. Pathological imaging groups may consequently be used as independent markers of pre-stroke cognitive impairment.

We found that women were older, had shorter education and higher mRS than men in this cohort. Despite this, more men had at least one pathological MRI finding, had more often lacunes and a pathological MTA. This difference remained after adjusting for age and gender. More men in our cohort had previous clinical stroke and hypertension. Also more men had AF, although borderline significance was reached (p=0.06). Increased cardiovascular and cerebrovascular risk factors in men, might be the reason why men who were on average younger showed similar odds for pre-stroke cognitive impairment as women who were on average older. When analyzing the imaging groups however, both men and women had increased OR for having a pathological GDS in the “cerebrovascular” and “mixed groups”. Even though numerically women had a higher OR for pathological GDS in the “mixed” and “cerebrovascular” groups, the difference was not significant. These observed odds were independent of gender by imaging interaction. Our study therefore does not show an increased risk of pre-stroke cognitive impairment in women, as was shown for pre-stroke dementia in the study by Pendlebury et al [7]. The combined effect of other risk factors in men and women such as higher education, which was independently associated with a reduced OR for pathological GDS, could have contributed to the numerically higher OR in women in the imaging groups. More studies are needed to disentangle the complex interactions of risk factors for pre-stroke cognitive impairment.

We chose a cut-off of GDS 2 as pathological in order to capture even participants with subtle memory complaints and subjective cognitive impairment (SCI) before stroke as these patients might also be prone to post-stroke cognitive impairment or dementia. The GDS [31] was designed for use in Alzheimer disease patients, but has also been validated for detection of vascular dementia [49]. Only 36% of the Nor-COAST patients with pathological MRI findings also had pathological scores on the GDS. The GDS is a scale that only provides an overview of larger stages of cognitive decline. Classification of cognitive impairment could probably better have been captured in a neuropsychological assessment, and perhaps it would have been easier to compare our results with other studies if a different scale had been used. In our study we needed a scale that could be used to retrospectively assess the patients without capturing the cognitive impairment of their acute stroke, and therefore chose the GDS scale.This may have led to underreporting of cognitive complaints in our cohort, leading to false negative results [50]. One reason for this could be that input on cognitive function came from relatives and some patients with a score of GDS 1, by us considered as normal cognition, could also have a mild form of cognitive impairment.

Many previous studies did not consider whether or not the scores were normal for the patient’s age. In “The Rotterdam Scan Study”, WMH were present in 95% of 1077 subjects (aged between 60-90) that were randomly sampled from the general population [18]. Compared to our finding of 38% pathological WMH in a stroke population, it seems there is an overlap between normal aging and SVD. We believe that cut-offs should be used whenever possible, to distinguish normal aging from pathological changes. Imaging findings of both SVD and neurodegeneration are frequent in stroke populations. They coexist often in the same individal and should therefore, as in the present study, be investigated together to determine the complete disease burden of a patient.

One advantage of visual scales is the opportunity to evaluate the visual scores of an individual as normal or pathological. The visual rating scales that we used are robust, validated and well suited both for the routine clinical setting as well as for research scans.Cut-offs help to interpret the visual rating scales and are practical tools in the decision-making process of clinicians [14]. Another strength of our study is the large number of participants included in a prospective multicenter study, with good quality brain MRIs. Another strength is that patients are not excluded based on their pre-stroke cognitive function. This reduces a possible selection bias towards patients with better cognitive function, although Aamodt (master student in our group, personal communication) showed that the patients who were not included in the MRI- part of the study were sicker than those who were included.

The use of visual rating scales can also be seen as a weakness as opposed to quantitative methods. Cut-offs for visual rating scales are important tools to interpret the visual rating, but cases that fall between categories are affected by some degree of uncertainty. Valuable information may get lost when grading subjects into a few categories, typically 0-3 or 0-4 in these rating scales. We will therefore continue to investigate the same population with volumetric methods. Research based cut-offs for the Fazekas scale are not described and one has to resort to cut-offs traditionally used in clinical practice [51]. Another weakness of the study is the lack of an age matched control group. This has been a decision in the study design that affects all work packages of NorCOAST, not only the imaging work package. Because of this limitation we cannot generalize our findings in this stroke cohort to a general ageing population.

## Conclusion

The majority of stroke patients in the Nor-COAST study had preexisting structural brain pathology. For the pathogenesis of pre-stroke cognitive impairment SVD seems to be more important than neurodegeneration. Chronic pathological changes may lead to impaired brain resilience [52]. Reduced brain resilience represents a risk factor for early- and late-onset post-stroke cognitive impairment and dementia [53]. This association makes imaging markers of chronic brain changes promising predictors for post-stroke cognitive impairment [53]. In future studies we will take our research on pre-stroke cognitive impairment and see how these brain changes are associated with early- and late onset poststroke cognitive impairment in the longitudinal follow-up of the same patient cohort. Future studies will reveal if there are gender differences in the risk of post stroke dementia.

## Data Availability

The current dataset cannot be made publicly available for ethical reasons, and public availability would compromise patient confidentiality and participant privacy. The study was conducted in humans and the dataset includes sensitive and personal information on individuals. A portion of data can be made available upon request to interested, qualified researchers provided that an agreement is made up. The minimal data set will enable replication of the reported study findings. Requests to access the datasets should be directed to [Mona K Beyer]

## Acknowledgements

The Nor-COAST collaboration group: Askim T., Aam S., Aaslund M.K., Aamodt E., Beyer M.K., Ellekjær H., Einstad M.S., Fure B., Gynnild M.A., Hamre C., Ihle-Hansen H., Knapskog A.B., Kummeneje C.S., Munthe-Kaas R., Næss H., Pendlebury S.T., Saltvedt I., Schellhorn T., Seljeseth Y., Thingstad P., Ursin M.

We would also like to thank all participants and their relatives for volunteering for this project. Thanks also to staff at all participating hospitals for providing access to scanners and images, and helping with all different practical tasks. Without you there would be no study. A special thank to Nina Sjøgren for helping with any thinkable task along the way. Thanks to Eva B Aamodt for language editing of the manuscript.

